# Gaps in Etiologic Classification of Chronic Kidney Disease in US Veterans: A Retrospective Cohort Study

**DOI:** 10.64898/2026.10.22.26365664

**Authors:** Julia J. Scialla, Kingsley Dwomoh, Benjamin Silverberg, Anisha Gerber, Wei Yu, Fei Heng, A. Taylor Kelley, Alfred K. Cheung, Guofen Yan

## Abstract

Many patients living with chronic kidney disease (CKD) may not have a clearly identified cause, which can be a barrier to targeted therapies. We assessed the assignment of CKD etiologies in the national Veterans Health Administration (VHA) electronic health record data, overall and by demographics. We identified regular VHA users aged 18 - 100 years with new-onset CKD based on persistent eGFR <60 ml/min/1.73m^2^ occurring 2005-2015. CKD etiologies were identified by administrative codes over 3 years before and after the incident CKD date. Etiologies were categorized as diabetic, hypertensive/vascular, obstructive/urologic, glomerular, cystic/congenital, and tubulointerstitial. CKD was considered recognized, but without etiology, if only a general CKD code was present. Logistic regression models assessed disparities by sex and race/ethnicity after adjustment for age, US Census region, and calendar year of incident CKD. Among 452,851 Veterans with CKD, 42.5% were assigned at least one etiology. Hypertensive/vascular (24.5%) was the most common. After adjustment, non-Hispanic Black Veterans and Hispanic Veterans were more likely to be assigned an etiologically-specific diagnosis than non-Hispanic White Veterans (OR 1.61 and 1.23, respectively, each p<0.001). Differences were greater at younger ages. Non-Hispanic Black Veterans were much more likely to be assigned a hypertensive/vascular etiology (OR 2.17; p<0.001), but both non-Hispanic Black and Hispanic Veterans were more likely than White veterans to be assigned diabetic and glomerular etiologies as well (each p<0.001). Women were much less likely to be assigned an etiology overall, and in all categories, compared to men (each p<0.001). Women were less likely to have CKD recognized even if etiology was not assigned. Overall, the assignment of an etiology for CKD is low among U.S. Veterans, particularly among women, possibly due to poor CKD recognition. Types of etiologies assigned differ by race/ethnicity, which may represent true etiologic difference or biased assignment.

## Introduction

Chronic kidney disease (CKD) affects approximately 14% of the US population and the burden is growing.^1–4^ CKD is defined by reduced glomerular filtration rate (GFR) or evidence of kidney damage persisting more than 90 days.^5^ However, CKD is a general term that encompasses many underlying etiologies of disease, including primary and secondary disorders. Different etiologies may have different natural history or treatment.^6^ This heterogeneity is recognized by international guidelines from Kidney Disease: Improving Global Outcomes (KDIGO), who recommend classifying CKD based on cause (C-stage), glomerular filtration rate (GFR or G-stage), and albuminuria (A-stage).^7^ While simple tools, such as estimated GFR equations and urine albumin-to-creatinine ratio (UACR), are available and recommended to guide classification in G and A-stages, guidance on determining cause is more limited. In a practice point, KDIGO guidelines suggest, “establish the cause of CKD using clinical context, personal and family history, social and environmental factors, medications, physical examination, laboratory measures, imaging, and genetic and pathologic diagnosis.”^7^ Currently available CKD toolkits do not emphasize this component of CKD evaluation,^8–13^ and underlying cause of CKD is often not identified.^14^ With a growing cadre of medications available to treat specific forms of CKD, this is a missed opportunity to provide disease-oriented care that may reduce risk of progression.

Prior studies focused on determining the cause for CKD have primarily studied specialty referral populations or focused on populations who may have had long-standing or advanced CKD. In one study within a specialty referral center in Germany, 20% of patients did not have a clear cause of CKD identified based on chart review at the time of enrollment into the study.^15^ In the US, reporting on the cause of CKD is typical in specific populations, such as those starting dialysis.^4^ There has been limited information on how often the cause of CKD is determined in a general adult CKD population, including in patients with new-onset CKD when kidney damage may still be reversible.

The national Veterans Health Administration (VHA), is the largest integrated health system in the US, providing an opportunity to study health services for chronic diseases, such as CKD, across often silo’ed settings, such as primary and specialty care or community and tertiary care. Our team has previously established a cohort of adult Veterans with regular VHA care utilization beginning from when they first develop CKD stage G3 or higher (defined as first GFR <60 ml/min/1.73m^2^ persisting for at least 90 days).^16^ Using this unique US cohort, our objective is to determine the proportion of adults with new-onset CKD who receive an etiologically-specific diagnosis within a timely manner. Our cohort avoids bias from specialty referral populations or prevalent and advanced CKD populations that have been assessed in most prior work. Due to widely described disparities in CKD outcomes and care according to race, ethnicity and sex,^16–20^ this study also evaluates how the assignment of etiologically-specific CKD diagnoses differs in these important demographic groups.

## Methods

### Data Source

This retrospective cohort study includes Veterans in the VHA, an integrated healthcare system spanning the US. Data sources include electronic health records (EHR) from the VHA Corporate Data Warehouse (CDW), VHA Vital Status File, and VHA annual utilization files, as well as administrative claims from the VHA/Centers for Medicare & Medicaid Services (CMS)/United States Renal Data System (USRDS) Data for Research, as previously described.^16^ The study was approved by the Institutional Review Board (IRB) at the University of Virginia and the joint IRB of the University of Utah and the Veterans Affairs Salt Lake City Healthcare System with a waiver of informed consent.

### Study Population and Cohort Creation

This study included adult US Veterans (18-100 years of age) with a new laboratory-based diagnosis of CKD stage G3 or higher (incident CKD G3+) between 2005-2015. Incident CKD G3+ was defined as the first occurrence of two outpatient eGFRs <60 ml/min/ 1.73m^2^ that were between 90 days and 18 months apart, with the index date corresponding to the second laboratory date in the pair. Because the goal of this study is to evaluate health care delivery, and the 2009 CKD-EPI equation was in use for most of the study period, we defined incident CKD G3+ using eGFR calculated from the 2009 CKD-EPI equation. To ensure that VHA healthcare data captured substantial portions of health care services, we only included veterans who were regular users of the VHA based on annual utilization files for 3 years before and after CKD. Veterans who died before completing 3 years of follow-up were excluded. Veterans who received international classification of disease (ICD-9 or ICD-10) diagnosis codes for CKD stage G3+ (i.e., 585.3-6; N18.3-6 or higher) before the index date were also excluded.

### Ascertainment of Etiologic Assignment

Specific etiologic diagnoses were ascertained by ICD-9 and ICD-10 billing codes in the 3 years prior to and following the index date of incident CKD stage G3+. We evaluated six etiologic domains including diabetic, hypertensive/vascular, obstructive/urologic, glomerular, cystic/congenital, and toxic/tubulointerstitial (**Supplemental Table 1**). The classification was designed by consensus between two nephrology experts (JJS and BS).

Patients who did not have an assigned etiology were further evaluated to see if CKD was recognized. CKD was considered recognized, but without an etiology, if there was only a general CKD code (ICD-9: 585.x; ICD-10: N18.x). Patients were classified as ‘CKD not recognized’ if they did not have either an etiologically-specific code or a general CKD code.

To allow descriptive summaries where more than one etiology code was found, mutually exclusive groupings were created using the logic depicted in **Figure 1**. The rationale for the classifications was to first select the etiology that would most meaningfully alter the course of CKD and therapy, while retaining some common combinations. The classification system was determined by consensus between two nephrology experts (JJS and BS).

**Figure 1:**
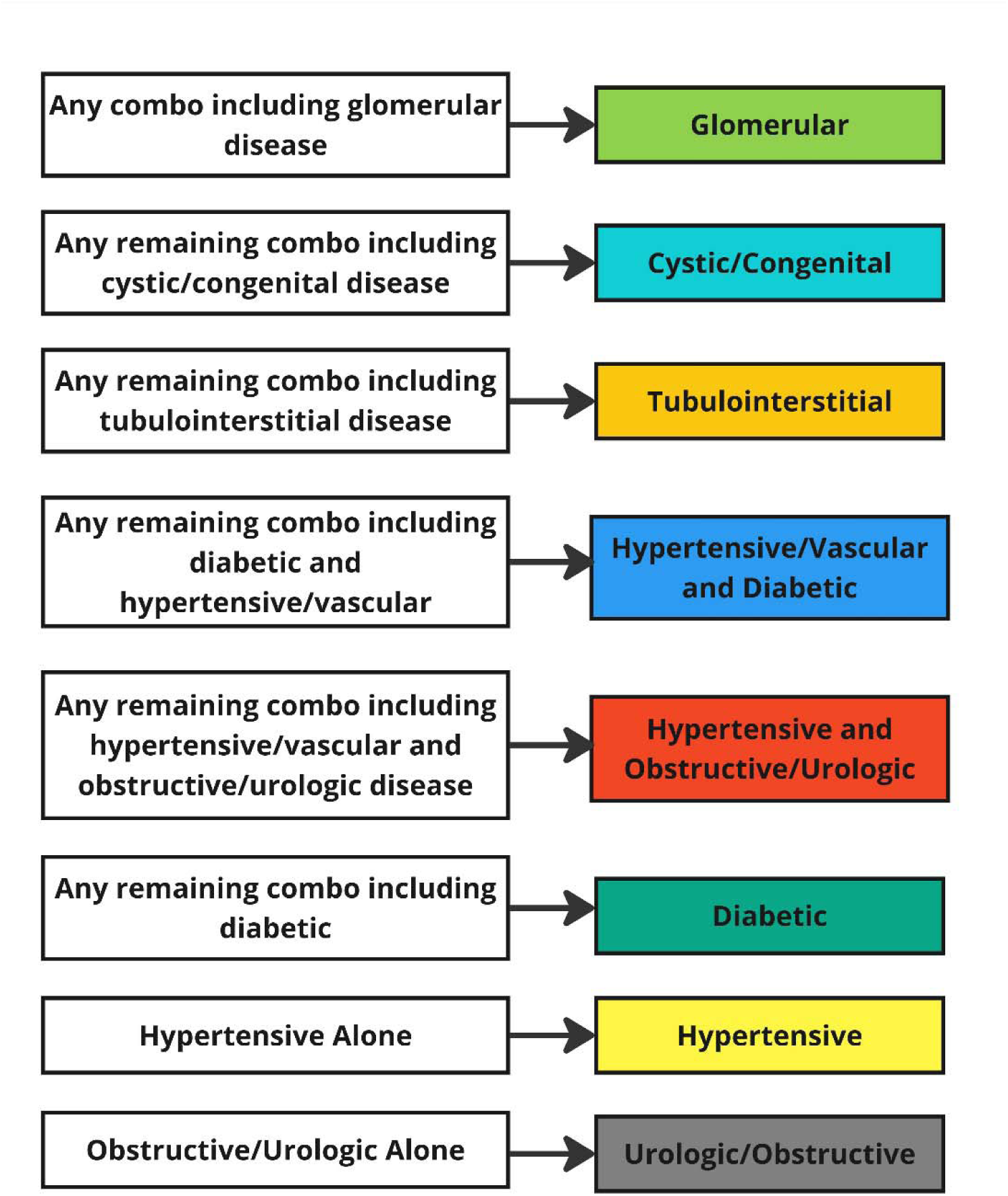
Hierarchical Assignment of Mutually Exclusive Etiologic Categories. Combinations of etiologies are possible therefore a hierarchy of common combinations was created focusing on the most important considerations for etiologies that may require unique therapy based on clinical rationale.

### Exposures and Covariates

Our analysis focuses on the assignment of etiologically-specific diagnoses overall and in subgroups defined by race/ethnicity (Non-Hispanic Black, Non-Hispanic White, Hispanic, Asian or Pacific Islander, Native American or Alaskan Native, Multiple or Other) and biological sex (male, female), each as listed in the VHA EHR. Simplified race/ethnicity groups were used for modeling of specific etiologies due to small sample size in some groups (Non-Hispanic Black, Non-Hispanic White, Hispanic, Other). Covariates included age (18-44; 45-64; 65-74; 75-100 years); the geographic region for VHA care using US Census designations, the year of the incident CKD G3+ index date, and selected comorbidities ascertained from administrative codes (**Supplemental Table 2**). Missing data in the cohort was minimal and restricted to individuals who were excluded for inadequate follow-up duration (5%).

### Statistical Analysis

The primary outcome was assignment of ≥1 administrative code for an etiologically-specific diagnosis of CKD. Secondary outcomes included assignment of ≥1 administrative code for a hypertensive/vascular etiology, assignment of ≥1 administrative code for a diabetic etiology, assignment of ≥1 administrative code for an obstructive/urologic etiology, and assignment of ≥1 administrative code for a glomerular etiology. We used logistic regression models to assess differences in assignment of any etiology and specific common etiologies by race/ethnicity and sex. We adjusted for age, geographic region, and year of incident CKD, as well as sex or race/ethnicity, as appropriate.

### Sensitivity Analyses

In sensitivity analyses, we: (1) additionally adjusted the main models for 13 common comorbidities including hypertension, diabetes, heart failure, coronary artery disease, cardiac dysrhythmia, other cardiac diseases, cerebrovascular accident/transient ischemic attack, peripheral vascular disease, chronic obstructive pulmonary disease, anemia, cancer, gastrointestinal bleeding disorders, and liver disease; (2) evaluated main modeling results stratified by age group; and, (3) evaluated main modeling results excluding patients who had an etiology assigned in the 3 years prior to the index date (n=358,292).

## Results

### Cohort Flow and Study Population

Among 732,707 Veterans who were identified by our research group to have incident CKD stage G3+ in the interval, 518,219 were identified as regular users of VHA healthcare services. Of these, 27,500 Veterans who died before completing the 3-year follow-up period and 37,868 Veterans with a diagnosis code for CKD stage G3+ prior to the index date were excluded. The final cohort included 452,851 patients.

The study population was predominantly male (96.8%); 52,653 (11.6%) were Non-Hispanic Black, 17,761 (3.9%) were Hispanic, and 368,397 (81.4%) were Non-Hispanic White. Mean age as 72 years with 0.5% 18-44 years old, 25.3% 45-64 years old, and the remainder 65 years or older. The participants were from all regions across the US with 41.9% from the South, 24.4% from the Midwest, 17.2% from the West, and 15.4% from the Northeast. Mean eGFR was 52.2 ml/min/1.73m^2^ at time of incident CKD G3+ and 9.8% had UACR ≥30 mg/g. Comorbidities including diabetes, hypertension and heart disease were common (**Table 1**).

**Table 1.** Characteristics of the Study Cohort at Incident Chronic Kidney Disease (CKD)

| **Characteristic**<br>**Mean ± SD or n (%)** | **Overall**<br>**(n=452,851)** |
| --- | --- |
| Age (years) | 71.7± 9.6 |
| 18-44 | 2,110 (0.5%) |
| 45-64 | 114,757 (25.3%) |
| 65-74 | 157,424 (34.8%) |
| 75-100 | 178,560 (39.4%) |
| Sex |  |
| Male | 438,414 (96.8%) |
| Female | 14,437 (3.2%) |
| Race/ethnicity |  |
| Non-Hispanic Black | 52,653 (11.6%) |
| Non-Hispanic White | 368,397 (81.4%) |
| Hispanic | 17,761 (3.9%) |
| Asian/Pacific Islander | 5,230 (1.2%) |
| Native American/Alaskan | 1,993 (0.4%) |
| Multiple or Other | 6,817 (1.5%) |
| Calendar Year of Incident CKD |  |
| 2005-2007 | 134,423 (29.7%) |
| 2008-2011 | 169,979 (37.5%) |
| 2012-2015 | 148,449 (32.8%) |
| US Census Region |  |
| Midwest | 110,684 (24.4%) |
| Northeast | 69,928 (15.4%) |
| South | 189,740 (41.9%) |
| West | 77,771 (17.2%) |
| US Territories | 4,728 (1.0%) |
| Comorbidities |  |
| Diabetes | 213,605 (47.2%) |
| Hypertension | 411,105 (90.8%) |
| Heart failure | 86,970 (19.2%) |
| Coronary artery disease | 203,175 (44.9%) |
| Cardiac dysrhythmia | 149,398 (33.0%) |
| Other cardiac diseases | 138,830 (30.7%) |
| CVA/TIA | 100,827 (22.3%) |
| Peripheral vascular disease | 124,623 (27.5%) |
| COPD | 129,699 (28.6%) |
| Anemia | 128,468 (28.4%) |
| Cancer | 99,694 (22.0%) |
| GI bleeding disorders | 54,660 (12.1%) |
| Liver disease | 17,097 (3.8%) |
| Estimated GFR (ml/min/1.73m<sup>2</sup>) | 52.2±7.0 |
| Urine albumin to creatinine ratio |  |
| <30 | 84,666 (18.7%) |
| 30-299 | 35,538 (7.8%) |
| ≥300 | 90,83 (2.0%) |
| Missing | 323,564 (71.5%) |

CVA/TIA, cerebrovascular accident/transient ischemic attack;
COPD, chronic obstructive pulmonary disorder

### CKD Etiologies in the Overall Population

Over the six-year total observation period, 42.5% of Veterans (n= 192,439) were assigned at least one etiologically-specific diagnosis, with 20.9% (n= 94,559) diagnosed during the 3-year baseline period, 35.9% (n= 162,714) during the 3-year follow-up, and 14.3% (n= 64,834) during both periods. Patients were more likely to receive an etiology in 2012-2015 (45.0%) than in 2005-2007 (40.9%), as were younger patients (53.9% for ages 18-44 years; 46.3% for ages 45-64 years; 42.8% for ages 65-74 years; and 39.7% for ages 75-100 years). The remaining 57.5% (n= 260,412) were not assigned any etiologically-specific diagnosis during the study window, although CKD was recognized based on a non-specific diagnosis code in an additional 10.5%.

The most common etiology assigned overall was hypertensive/vascular (24.5%; n= 111,145), followed by obstructive/urologic (18.4%; n= 83,488), and diabetic (11.1%; n= 50,170). Less frequent causes included glomerular disease (5.0%), cystic/congenital etiologies (4.9%), and toxic/tubulointerstitial etiologies (0.4%; **Supplemental Table 3**).

### Association of Race/Ethnicity with Etiologic Assignment

Non-Hispanic Black Veterans had the highest proportion with an etiology assigned (54.3%), followed by Hispanic Veterans (46.7%). CKD was recognized without an etiology in 13.7% of Non-Hispanic Black Veterans compared to 11.2% of Hispanic Veterans, 10.9% of Veterans of other races, and 10.0% of Non-Hispanic White Veterans. CKD was not recognized in a plurality of Non-Hispanic White Veterans (49.4%) and Veterans of other races (46.0%). CKD was more likely to be recognized than not in Hispanic Veterans and non-Hispanic Black Veterans, with CKD not recognized in 42.1%, and 32.0%, respectively (**Supplemental Figure 1**).

In non-Hispanic Black veterans, hypertensive/vascular alone (19.1%) was the most prevalent etiology, followed by glomerular diseases (8.5%) and hypertensive/vascular and diabetic (7.6%). In Hispanic Veterans, hypertensive/vascular alone (10.3%), obstructive/urologic diseases alone (9.4%), and glomerular disease (7.5%) accounted for the majority of etiologies. 40.6% of Non-Hispanic White Veterans had etiologies distributed across hypertensive/vascular alone (10.8%), obstructive/urologic diseases alone (10.6%), and cystic/congenital (4.5%) as top causes (**Figure 2A**).

**Figure 2.**
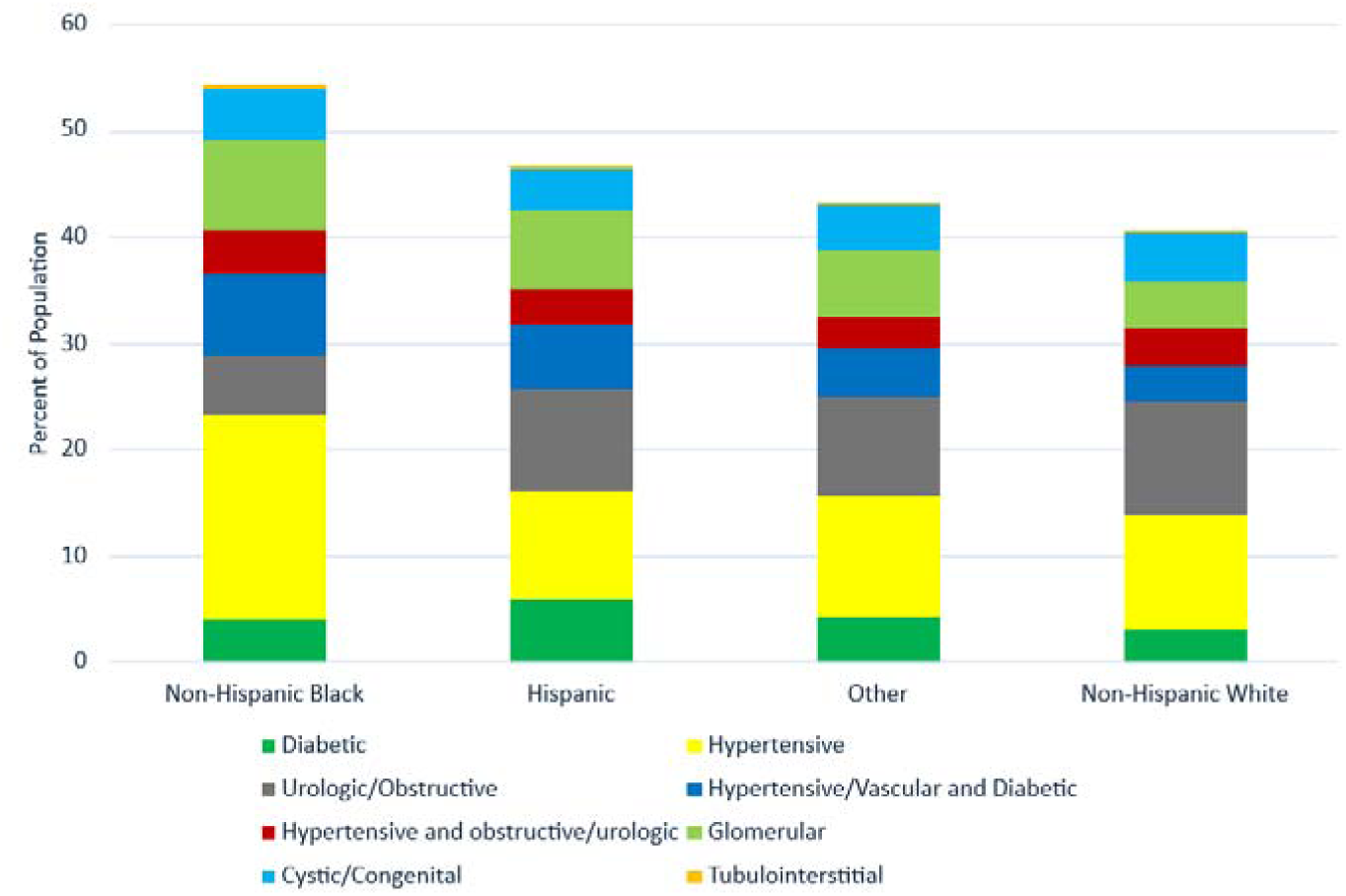
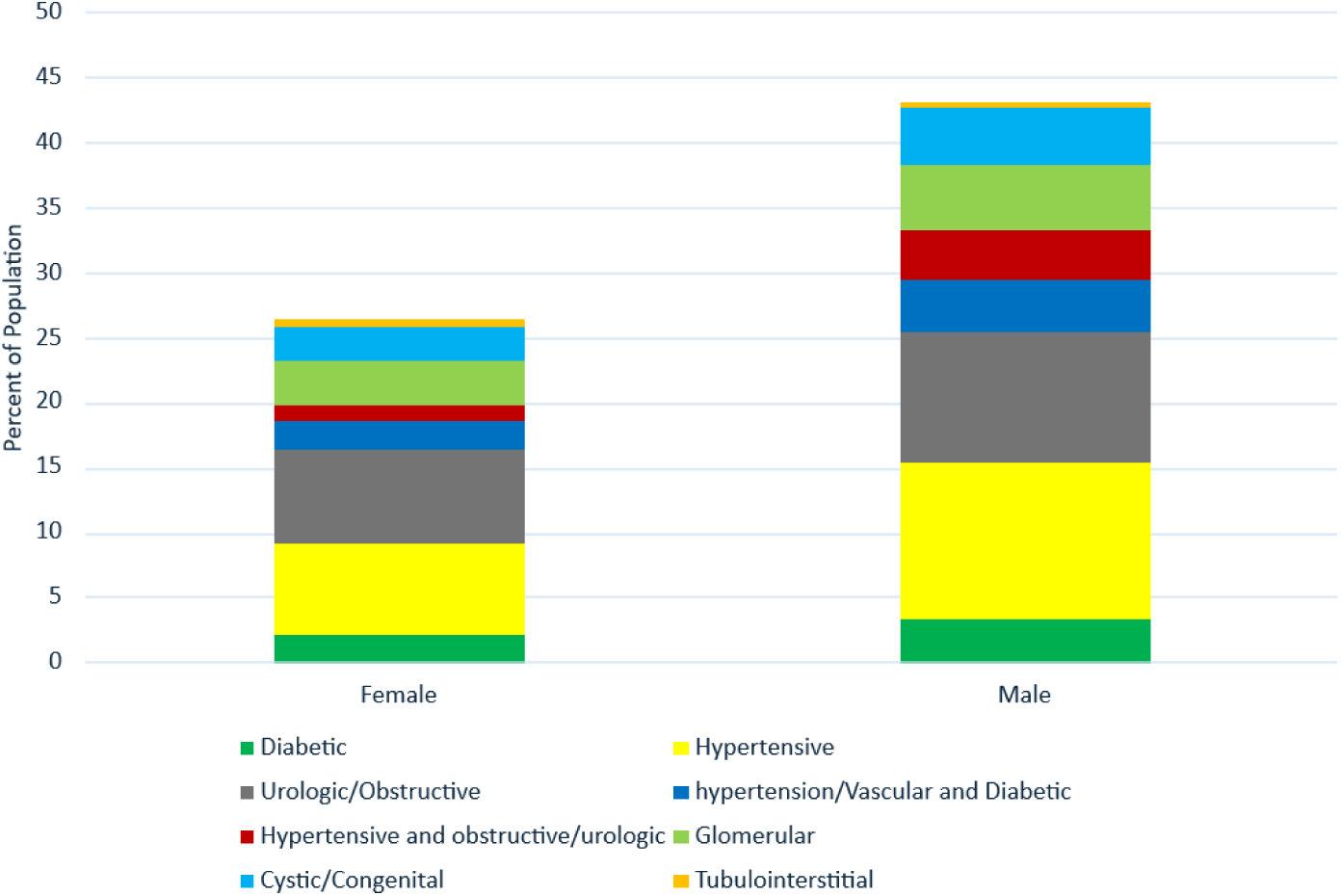
Etiologies Assigned Over Full Follow Up by Race (A) and Sex (B). Assigned etiologies are based on administrative codes over the 3 years prior and 3 years after the index incident CKD G3+ date. Colors within bars represent the proportion of the population with the etiologic category based on the mutually exclusive groupings in Figure 1. The total height of the bar represents the overall proportion of the population that received an etiologically-precise diagnosis.

Next, we built multivariable models of the odds of assigning an etiology after adjustment for age, sex, US Census region, and calendar year of incident CKD. Odds of assigning any etiology was higher for Non-Hispanic Black compared to Non-Hispanic White Veterans (OR 1.61; 95%CI 1.58-1.64; **Table 2**). Hispanic Veterans also had higher odds (OR 1.23; 95%CI 1.19-1.27), as did Asian/Pacific Islanders (p=0.03) and Native American/Alaska Natives (p=0.01).

**Table 2.** Odds of Receiving any Etiologic Assignment by Race/Ethnicity and Sex.

|  | Odds Ratio of Getting Etiology at Baseline or Follow-Up<br>N=452,851 |  | Odds Ratio of Etiology over Follow-Up Only<br>N=358,292 |  |
| --- | --- | --- | --- | --- |
|  | Adjusted OR (95% CI) | p-value | Adjusted OR (95% CI) | p-value |
| ***Race/Ethnicity*** |  |  |  |  |
| Non-Hispanic Black | 1.61 (1.58-1.64) | <0.001 | 1.66 (1.62-1.70) | <0.001 |
| Non-Hispanic White | Ref |  | Ref |  |
| Hispanic | 1.23 (1.19-1.27) | <0.001 | 1.25 (1.20-1.30) | <0.001 |
| Asian/Pacific Islander | 1.06 (1.01-1.13) | 0.03 | 1.09 (1.02-1.17) | 0.01 |
| Native American/<br>Alaskan | 1.12 (1.02-1.23) | 0.01 | 1.09 (0.97-1.22) | 0.1 |
| Multiple or Other | 1.08 (1.03-1.14) | 0.001 | 1.09 (1.03-1.16) | 0.006 |
| ***Biological Sex*** |  |  |  |  |
| Male | Ref |  | Ref |  |
| Female | 0.42 (0.40-0.43) | <0.001 | 0.44 (0.42-0.46) | <0.001 |

\*Adjusted for age, US Census region, calendar year at incident CKD, and sex or race/ethnicity, as relevant

Higher odds for Non-Hispanic Black Veterans were driven by higher assignment of hypertensive/vascular, diabetic and glomerular etiologies, and lower assignment of obstructive etiologies (**Table 3**). Results were similar when comparing Hispanic Veterans to Non-Hispanic White Veterans, with higher odds of assigning hypertensive/vascular, diabetic and glomerular etiologies, although enriched assignment of hypertensive/vascular etiology was less pronounced compared to enrichment in Non-Hispanic Black Veterans. (**Table 3**).

**Table 3.** Adjusted Odds of Receiving Specific Etiologic Assignments Over Full Follow-Up by Race/Ethnicity and Sex.

|  | Odds Ratio of Assigning Hypertensive/Vascular Etiology |  | Odds Ratio of Assigning Diabetic Etiology |  | Odds Ratio of Assigning Urologic/Obstructive Etiology |  | Odds Ratio of Assigning Glomerular Disease Etiology |  |
| --- | --- | --- | --- | --- | --- | --- | --- | --- |
|  | Adjusted OR (95% CI) | p-value | Adjusted OR (95% CI) | p-value | Adjusted OR (95% CI) | p-value | Adjusted OR (95% CI) | p-value |
| ***Race/Ethnicity*** |  |  |  |  |  |  |  |  |
| Non-Hispanic Black | 2.17 (2.13-2.21) | <0.001 | 1.70 (1.66-1.75) | <0.001 | 0.76 (0.74-0.78) | <0.001 | 1.67 (1.61-1.73) | <0.001 |
| Non-Hispanic White | Ref |  | Ref |  | Ref |  | Ref |  |
| Hispanic | 1.32 (1.27-1.37) | <0.001 | 1.80 (1.72-1.88) | <0.001 | 0.93 (0.89-0.97) | 0.001 | 1.73 (1.63-1.85) | <0.001 |
| Other | 1.10 (1.05-1.14) | <0.001 | 1.36 (1.29-1.43) | <0.001 | 0.90 (0.86-0.94) | <0.001 | 1.33 (1.24-1.43) | <0.001 |
| ***Biological Sex*** |  |  |  |  |  |  |  |  |
| Male | Ref |  | Ref |  | Ref |  | Ref |  |
| Female | 0.38 (0.36-0.40) | <0.001 | 0.36 (0.34-0.39) | <0.001 | 0.55 (0.52-0.58) | <0.001 | 0.44 (0.40-0.48) | <0.001 |

\*Adjusted for age, US Census region, calendar year at incident CKD, and sex or race/ethnicity, as relevant

In sensitivity analyses, differential odds of etiologic assignment by race/ethnicity remained after additional adjustment for comorbidities (**Supplemental Table 4**), or if excluding patients with an etiology assigned at baseline (**Table 2**). Disparities in etiologic assignment by race were more pronounced in younger, compared to older, adults (**Table 4**).

**Table 4.**
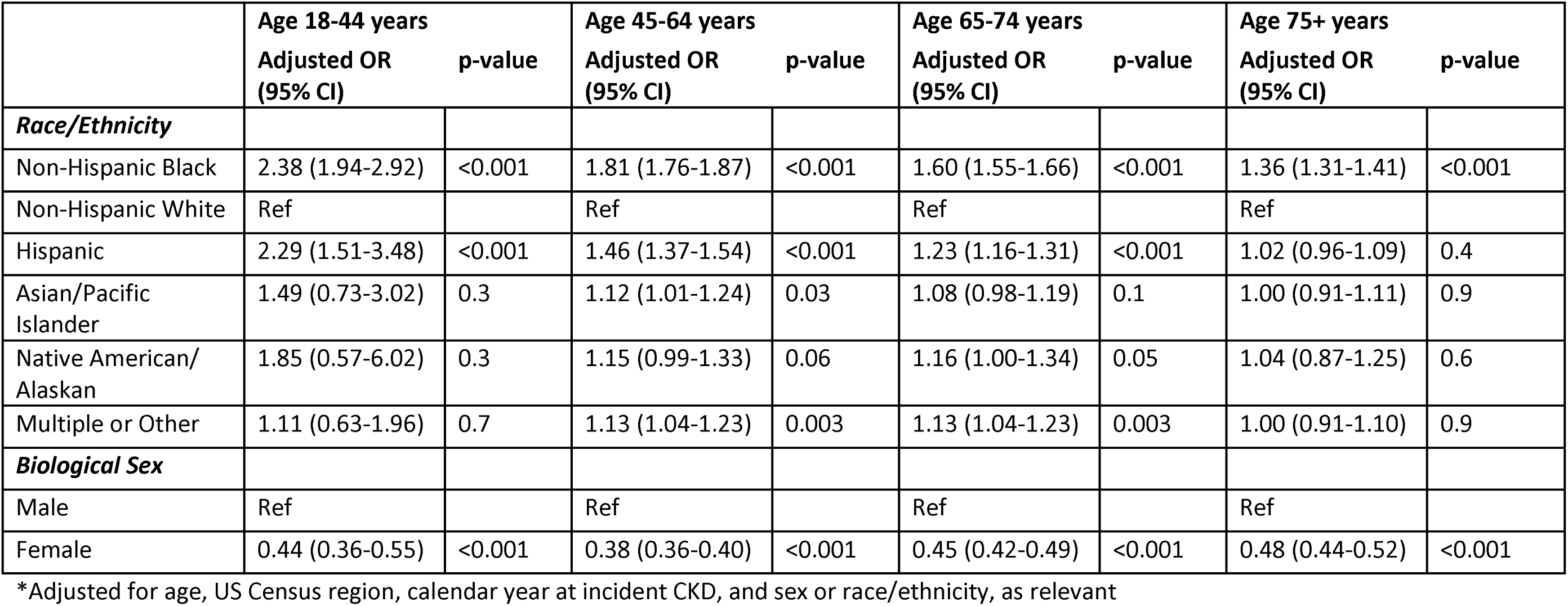
Age-Stratified, Adjusted Odds of Receiving Specific Etiologic Assignments Over Full Follow-Up by Race/Ethnicity and Sex.

|  | **Age 18-44 years** |  | **Age 45-64 years** |  | **Age 65-74 years** |  | **Age 75+ years** |  |
| --- | --- | --- | --- | --- | --- | --- | --- | --- |
|  | **Adjusted OR**<br>**(95% CI)** | **p-value** | **Adjusted OR**<br>**(95% CI)** | **p-value** | **Adjusted OR**<br>**(95% CI)** | **p-value** | **Adjusted OR**<br>**(95% CI)** | **p-value** |
| ***Race/Ethnicity*** |  |  |  |  |  |  |  |  |
| Non-Hispanic Black | 2.38 (1.94-2.92) | <0.001 | 1.81 (1.76-1.87) | <0.001 | 1.60 (1.55-1.66) | <0.001 | 1.36 (1.31-1.41) | <0.001 |
| Non-Hispanic White | Ref |  | Ref |  | Ref |  | Ref |  |
| Hispanic | 2.29 (1.51-3.48) | <0.001 | 1.46 (1.37-1.54) | <0.001 | 1.23 (1.16-1.31) | <0.001 | 1.02 (0.96-1.09) | 0.4 |
| Asian/Pacific<br>Islander | 1.49 (0.73-3.02) | 0.3 | 1.12 (1.01-1.24) | 0.03 | 1.08 (0.98-1.19) | 0.1 | 1.00 (0.91-1.11) | 0.9 |
| Native American/<br>Alaskan | 1.85 (0.57-6.02) | 0.3 | 1.15 (0.99-1.33) | 0.06 | 1.16 (1.00-1.34) | 0.05 | 1.04 (0.87-1.25) | 0.6 |
| Multiple or Other | 1.11 (0.63-1.96) | 0.7 | 1.13 (1.04-1.23) | 0.003 | 1.13 (1.04-1.23) | 0.003 | 1.00 (0.91-1.10) | 0.9 |
| ***Biological Sex*** |  |  |  |  |  |  |  |  |
| Male | Ref |  | Ref |  | Ref |  | Ref |  |
| Female | 0.44 (0.36-0.55) | <0.001 | 0.38 (0.36-0.40) | <0.001 | 0.45 (0.42-0.49) | <0.001 | 0.48 (0.44-0.52) | <0.001 |

\*Adjusted for age, US Census region, calendar year at incident CKD, and sex or race/ethnicity, as relevant

### Association of Biological Sex with Assignment of Etiology

Approximately 43.0% of male Veterans had an etiology assigned, in contrast to 26.3% of female Veterans. Among female Veterans without an assigned etiology (n=10,634), 1,285 (8.9% of the total population) had CKD recognition, while the majority of the total female population (64.8%) had no CKD recognition. In comparison, 46,211 (10.5% of total male population) male Veterans without an assigned etiology had CKD recognition, and 46.4% of the total male population had no CKD recognition (**Supplemental Figure 2**).

In male Veterans, hypertensive/vascular alone was the most commonly assigned etiology (12.0%), followed by obstructive/urologic diseases alone (10.0%), and glomerular diseases (5.0%). In female Veterans obstructive/urologic diseases alone (7.3%), hypertensive/vascular alone (6.9%), and glomerular disease (3.3%) accounted for the majority of identified cases (**Figure 2B**).

Female Veterans had lower odds of etiologic assignment in adjusted models (OR 0.42; 95%CI 0.40-0.43; p<0.001; **Table 2**). Female veterans had lower rates of being assigned hypertensive/vascular, diabetic, obstructive/urologic, and glomerular etiologies compared to their male counterparts (**Table 3**).

In sensitivity analyses, differential odds of etiologic assignment were similar after additional adjustment for comorbidities (**Supplemental Table 4**), or if excluding patients with an etiology assigned at baseline (**Table 2**). Disparities in etiologic assignment by sex were similar across age strata (**Table 4**).

## Discussion

In this study using an incident CKD cohort from the national VHA Health System, we found a relatively low rate of assigning timely, etiologically-specific CKD diagnoses, such as hypertensive/vascular, diabetic, obstructive/urologic, cystic/congenital, glomerular, or toxic/tubulointerstitial diagnoses, according to administrative billing codes. For a large fraction of patients without an etiology assigned, CKD was also not recognized by a non-specific code, making it a potentially major factor in limiting etiologic assessment. The rates of CKD recognition and etiologically-specific diagnoses were particularly low in female Veterans, consistent with other studies demonstrating disparities for women with other aspects of CKD evaluation and management.^21–25^ Non-Hispanic Black and Hispanic Veterans had higher rates of etiologically-specific diagnoses, particularly at younger age, but these were dominated by hypertensive/vascular and diabetic etiologies. It is unclear if the preponderance of hypertensive and diabetic diagnoses in this group reflects true differences in disease or a biased evaluation with assignment of diagnoses that may be convenient or assumed, rather than accurate.^26^ In light of pervasive disparities in CKD outcomes and management for non-Hispanic Black and Hispanic patients compared to White patients,^16,19,25,27,28^ and women compared to men,^21–25^ further research into the CKD evaluation is needed to ensure that an appropriate diagnostic evaluation is performed for all individuals with CKD.

Determining the etiology of CKD is supported in international staging systems.^7^ A recent consensus statement from the European Renal Association emphasizes why etiologic resolution may be particularly important:^26^ namely, as we enter an age of molecular diagnostics and treatment, unsolved cases of CKD may be at a disadvantage. Studies show that a substantial fraction of CKD of truly unknown etiology can be resolved by genetic diagnosis.^29^ However, this refers to cases in which a medically appropriate, but comprehensive evaluation has already been completed without a clear cause, what the consensus group refers to as CKDx.^26^ A greater focus on the evaluation for etiologically-specific diagnoses in CKD could help to usher in advances from molecular medicine.

CKD of different etiologies often have different natural history that could impact prognosis, referral, monitoring, and treatment. A recent analysis within the German CKD Cohort found much higher rates of GFR decline among CKD patients with primary glomerular or diabetic kidney disease compared to other forms.^30^ The disease type was a stronger predictor than starting eGFR and UACR, which dominate the current staging system. A recent study of patients with IgA nephropathy demonstrated dramatically increased rates of progression and kidney failure compared to the natural history of other non-glomerular forms of CKD.^31^ The different behavior of specific forms of CKD underscores the need for more research on how to help clinicians clearly establish a cause of CKD in most cases. This precision approach may be in tension with the growing emphasis on Cardiovascular-Kidney-Metabolic (CKM) Syndrome that conceptualizes CKD within a framework of metabolic risks related to excess adiposity, hypertension and other lifestyle factors that are common risk factors for CKD, but not exclusive causes.^32^ As CKM training enters primary care, it will be important to ensure that clinicians understand when a different, more specific, disease paradigm may better apply (e.g., obstructive, cystic, or glomerular disease).

The rates of etiologically-specific diagnoses were particularly low among female Veterans. Women with CKD have been shown to have lower rates of transplant referral,^21^ treatment with disease-modifying therapy, and other diagnostic evaluations, such as evaluation of hematuria.^22,28^ It is possible that these disparities reflect poor recognition of CKD, which may be present even when serum creatinine concentrations may appear normal or near-normal due to differences in creatinine production by sex.^33^ Alternatively, other signs such as hematuria or proteinuria may be ignored, assuming these are related to menstruation or urinary tract infections that are more common in women than men. Another possibility is that this may be related to a lower expected risk in women,^24^ or cognitive bias. Whatever the reason, lower rates of etiologic assignment may limit appropriate care for women with CKD and further study is needed to understand the reasons and remedies for these observations. We acknowledge that women represent a significant minority in the VHA Health System, but due to our large sample size there were a large number of women represented, and our power was adequate for these important comparisons.

Differences in etiologic assignment were also noted by race and ethnicity. Non-Hispanic Black patients and Hispanic patients were more likely to be assigned an etiology compared to White patients overall, especially at younger age. However, some of the largest adjusted differences were for hypertensive/vascular and diabetic etiologies. Hypertensive nephropathy, in particular, has been called into question as a common etiology and instead may often be a consequence of kidney disease or a feature of genetic glomerular diseases, such as APOL-1 associated nephropathy.^26,34–37^ In this study, hypertensive/vascular causes were particularly enriched among Black Veterans, a phenomenon that has been described for decades, including contemporary reports of the US Renal Data System.^4,38^ Case vignette studies have demonstrated that identifying a patient’s race as Black led to nearly twice the rate of attributing kidney disease to ‘hypertensive’ compared to identifying a patient’s race as White, suggesting bias in disease attribution.^39^

Similarly, diabetes can be a cause or comorbidity of CKD,^40^ and, in our study, was also enriched as an etiologic diagnosis in Non-Hispanic Black and Hispanic Veterans compared to White Veterans. A recent study of genetic testing in a general CKD cohort demonstrated a diagnostic yield for 14% in those with suspected hypertensive nephropathy and 11% in those with suspected diabetic nephropathy.^41^ These results suggest possible erroneous attribution of CKD to diabetes and hypertension without a more thorough evaluation. Our study is not able to determine the appropriate evaluation or the accuracy of the assigned CKD etiology, but does highlight differences in attribution by race and ethnicity that should be further studied. Our team’s prior work demonstrates that Black Veterans develop CKD, on average, 8 years younger than White Veterans,^42^ and that younger Black Veterans also have higher rates of kidney function decline than White Veterans:^43^ factors that may be hallmarks of genetic disorders.^44,45^ With these prior results in mind, it will be critical to determine if inaccurate or abbreviated evaluation in young Black Veterans contributes to these disparities.

This study has many strengths including a large population with rigorous construction to ensure anchoring of analyses in new-onset CKD. The VHA is the largest integrated healthcare system in the US, with good geographical and demographic representation. With a focus on patients with regular VHA utilization, we ensure near comprehensive capture of healthcare activities that should minimize risks of missing assessment or diagnoses assigned outside of the VHA. We assessed a broad range of administrative codes to define etiologically-specific diagnoses.

We also recognize some important limitations. Administrative diagnoses can be inaccurate. They may not fully capture the extent of diagnoses that are recognized by clinicians, but not billed. Diabetic and tubulointerstitial etiologies may be particularly limited due to changes in coding options between ICD-9 and ICD-10. Despite this, demographic comparisons across patient groups and across diagnosis types would be expected to be robust to these limitations. Here we focused on demographic factors. Other important factors could include academic affiliation, provider types, regional variability and others, and should be assessed in future studies. The data that we used for this analysis are from 2005-2018 and may not fully represent a more contemporary dataset. We did not utilize a more contemporary era to avoid disruption due to the COVID-19 pandemic that would impact the required 6-year participation period for individuals with new-onset CKD after 2015. Finally, our analysis focused on assignment of etiologically-specific diagnoses. In a large fraction of cases in which we did not detect an etiologically-specific diagnosis, we also found that there was no general administrative code for CKD, suggesting that CKD may not have been recognized. Future studies should assess a prospective sequence of evaluation that would progress from recognition to risk stratification, appropriate testing, followed by etiologically-specific diagnoses.

In conclusion, in the US non-Hispanic Black and Hispanic Veterans compared to non-Hispanic White Veterans, and male Veterans compared to female Veterans are more likely to receive etiologically-specific diagnostic codes. Non-Hispanic Black and Hispanic Veterans had higher rates of being assigned hypertensive/vascular and diabetic etiologies that may reflect bias in diagnostic evaluation worthy of further investigation. These results should be confirmed in other health systems to ensure generalizability. Guidelines for a structured CKD evaluation could help reduce differences in assignment of etiologically-specific CKD diagnoses by race/ethnicity and sex.

## Data Availability

Per the terms of the data use agreement with the Veterans Health Administration, data cannot be made publicly available.

## Acknowledgements

This study was supported in part by R01DK112008 from the National Institutes of Diabetes and Digestive and Kidney Diseases (NIDDK). The funding agency had no role in study design, data collection and analysis, decision to publish, or preparation of the manuscript. VA/CMS data was provided by the Department of Veterans Affairs, Veterans Health Administration, Office of Research and Development, Health Services Research and Development, VA Information Resource Center. Support for accessing VA national databases and computations was provided by the Veterans Informatics and Computing Infrastructure (VINCI) team in Salt Lake City, Utah. The contents of this work are solely the responsibility of the authors and do not necessarily represent the official views of the NIDDK, National Institutes of Health, or the Veterans Health Administration. This work was previously presented in part in abstract form at American Society of Nephrology’s Kidney Week 2023.

