## Supplemental Material for "Gaps in Etiologic Classification of Chronic Kidney Disease in US Veterans: A Retrospective Cohort Study"

**Supplemental Table 1. Administrative Definition of Etiologically-Precise CKD Diagnoses**

| **Category** | **Example Diagnoses** | **ICD-9 Codes** | **ICD-10 Codes** |
| --- | --- | --- | --- |
| **Glomerular** | Membranous nephropathy | 283.11, 580-583 | D59.3, N00-N08, M32.14 |
| **Toxic/Tubulo-interstitial** | Sjogren’s syndrome with nephritis | n/a* | M32.15, M35.04, N11.x, N12, N14.x, N15.x, N16 |
| **Hypertensive/ Vascular** | Hypertensive Nephropathy; Heart Failure; Renal Artery Stenosis | 403, 404, 405.01, 405.11, 405.91, 593.81, 642.4x, 642.5x, 642.6x, 642.7x | I12, I13, I15.0, N28.0, O11, 014, 015 |
| **Cystic/**  **Congenital** | Polycystic Kidney Disease; Congenital Renal Agenesis | 753.0, 753.1x, | Q60, Q61.00, Q61.02, Q61.1x, Q61.2, Q61.3, Q61.4, Q61.5, Q61.8, Q61.9 |
| **Diabetic** | Diabetic Nephropathy | 249.4x, 250.4x | E08.2x, E09.2x, E10.2x, E11.2x, E13.2x |
| **Obstructive/**  **Urologic** | Nephrolithiasis; Nephrectomy; Hydronephrosis | 591, 592.x, 593.3, 593.4, 593.5, 593.7x, 593.89, 599.6x, 753.2x, V10.52, V45.73 | N13.X, N13.0, N13.1, N13.2, N13.3x, N13.4, N13.7x, N13.8, N13.9, N20.x, Q62.x, Z85.528, Z90.5 |
| *Chronic tubulointerstitial disease introduced only in ICD-10 | | | |

**Supplemental Table 2. Administrative Definition of Comorbidities**

| **Comorbidity** | **ICD-9-CM codes** | **ICD-10-CM codes** |
| --- | --- | --- |
| **Anemia** | 280-285 | D50.0-D64.9 |
| **Cancer** | 140-172; 174-208; 230-231; 233-234 | C00.0-C43.9; C45.0-C75.9; C76.0-D03.9; D05.00-D09.9 |
| **Cardiac, other** | 420-424; 429; 785.0-785.3; V42.2; V43.3 | A18.84; I23.0-I23.8; I25.10; I30.0-I39; I51.0- I52; I97.0-I97.191; M32.11; M32.12; R00.0; R00.2-R01.2; Z95.2-Z95.4 |
| **Cerebrovascular accident (CVA) / transient ischemic attack (TIA)** | 430-438 | G45.0-G45.2; G45.4-G46.8; I60.00-I66.9; I67.1; I67.2; I67.4-I67.82; I67.841-I69.998 |
| **Chronic obstructive pulmonary disorder (COPD)** | 491-494; 496; 510 | J41.0-J47.9; J86.0; J86.9 |
| **Coronary artery disease** | 410-414; V45.81; V45.82 | I12.00-I22.9; I24.0-I25.9; Z95.1; Z95.5; Z98.61 |
| **Diabetes mellitus** | 250; 357.2; 362.0; 366.41 | E08.311-E08.36; E08.40; E08.42; E09.311- E09.36; E09.40; E09.42; E10.10-E13.9 |
| **Cardiac dysrhythmia** | 426-427; V45.0; V53.3 | I44.0-I49.9; R00.1; Z45.0-Z45.09; Z95.0; Z95.810; Z95.818; Z95.9 |
| **Gastrointestinal bleeding disorders (GI)** | 456.0-456.2; 530.7; 531-534; 569.84-  569.85; 578 | I85.00-I85.11; K22.6; K25.0-K28.9; K55.20; K55.21; K56.60; K92.0-K92.2 |
| **Heart failure** | 398.91; 402.01, 402.11, 402.91; 404.01,  404.03, 404.11, 404.13, 404.91, 404.93;  422; 425; 428; V42.1 | A18.84; I09.81; I11.0; I13.0; I13.2; I40.0-I43; I50.1-I50.9; Z48.21; Z48.280; Z94.1; Z94.3 |
| **Hypertension** | 362.11; 401-405; 437.2 | H35.031-H35.039, I10-I13.2, I15.0-I15.9, I67.4, N26.2 |
| **Liver disease** | 570-571;572.1, 572.4; 573.1-573.3; V42.7 | B25.1; K70.0-K72.01; K73.0-K74.69; K77; Z48.23; Z94.4 |
| **Peripheral vascular disease (PVD)** | 440-444; 447; 451-453; 557 | E08.51; E08.52; E09.51; E09.52; E10.51; E10.52; E11.51; E11.52; E13.51; E13.52; I67.0; I70.0-I74.9; I77.0-I77.9; I79.0-I82.91;  K55.0; K55.1; K55.8; K55.9; M31.8; M31.9 |

**Supplement Table 3: Rates of specific etiologies in the overall cohort**

| **Etiology** | **Frequency** | **Percent** |
| --- | --- | --- |
| Hypertensive/Vascular | 111,145 | 24.5% |
| Obstructive/Urologic | 83,488 | 18.4% |
| Diabetic | 50,170 | 11.1% |
| Glomerular | 22,562 | 5.0% |
| Cystic/Congenital | 22,023 | 4.9% |
| Toxic/Tubulointerstitial | 1,657 | 0.4% |

**Supplemental Table 4. Odds of Receiving any Etiologic Assignment by Race/Ethnicity and Sex, Additionally Adjusted for Comorbidities**

|  | **Odds Ratio of Getting Etiology at Baseline or Follow-Up**  **N=452,851** | | **Odds Ratio of Etiology over Follow-Up Only**  **N=358,292** | |
| --- | --- | --- | --- | --- |
|  | **Adjusted OR (95% CI)** | **p-value** | **Adjusted OR (95% CI)** | **p-value** |
| ***Race/Ethnicity*** |  |  |  |  |
| Non-Hispanic Black | 1.56 (1.53-1.59) | <0.001 | 1.63 (1.59-1.67) | <0.001 |
| Non-Hispanic White | Ref |  | Ref |  |
| Hispanic | 1.17 (1.13-1.21) | <0.001 | 1.18 (1.13-1.23) | <0.001 |
| Asian/Pacific Islander | 1.09 (1.03-1.15) | 0.005 | 1.09 (1.02-1.17) | 0.01 |
| Native American/  Alaskan | 1.11 (1.01-1.22) | 0.03 | 1.09 (0.97-1.22) | 0.1 |
| Multiple or Other | 1.04 (0.98-1.09) | 0.2 | 1.05 (0.99-1.12) | 0.1 |
| ***Biological Sex*** |  |  |  |  |
| Male | Ref |  | Ref |  |
| Female | 0.47 (0.45-0.48) | <0.001 | 0.49 (0.46-0.51) | <0.001 |

*Adjusted for age, US Census region, calendar year at incident CKD, 13 comorbidities, and sex or race/ethnicity, as relevant

**Supplement Figure 1. Proportion of the study cohort with CKD etiology, CKD recognized with no etiology, and CKD not recognized by race/ethnicity.** CKD was considered recognized if the patient had an administrative code for an etiologically-specific diagnosis or a general non-specific CKD code. Patients with neither code type in the interval were considered not recognized, and patients with only a general non-specific CKD code were recognized, but without an etiology.

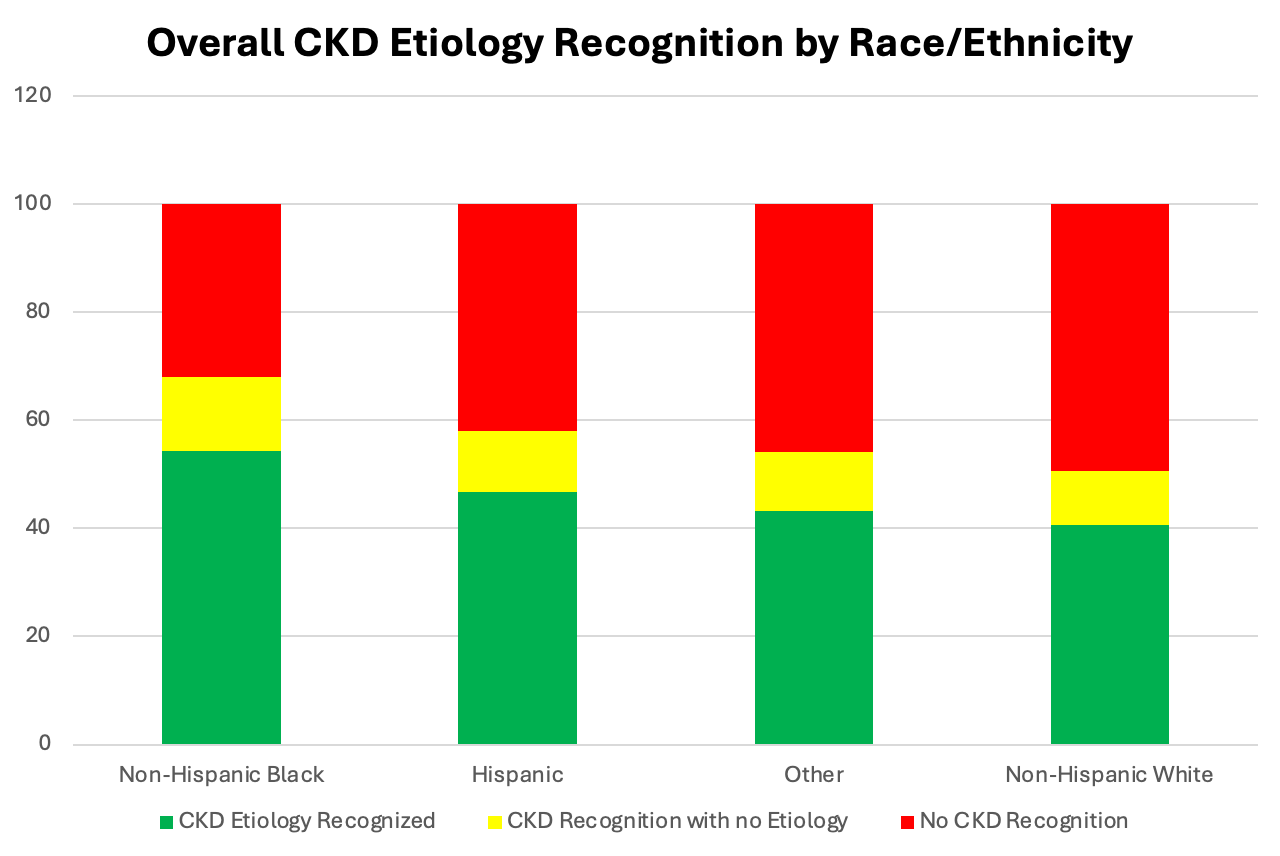

**Supplement Figure 2. Proportion of the study cohort with CKD etiology, CKD recognized with no etiology, and CKD not recognized by sex.** CKD was considered recognized if the patient had an administrative code for an etiologically-specific diagnosis or a general non-specific CKD code. Patients with neither code type in the interval were considered not recognized, and patients with only a general non-specific CKD code were recognized, but without an etiology.

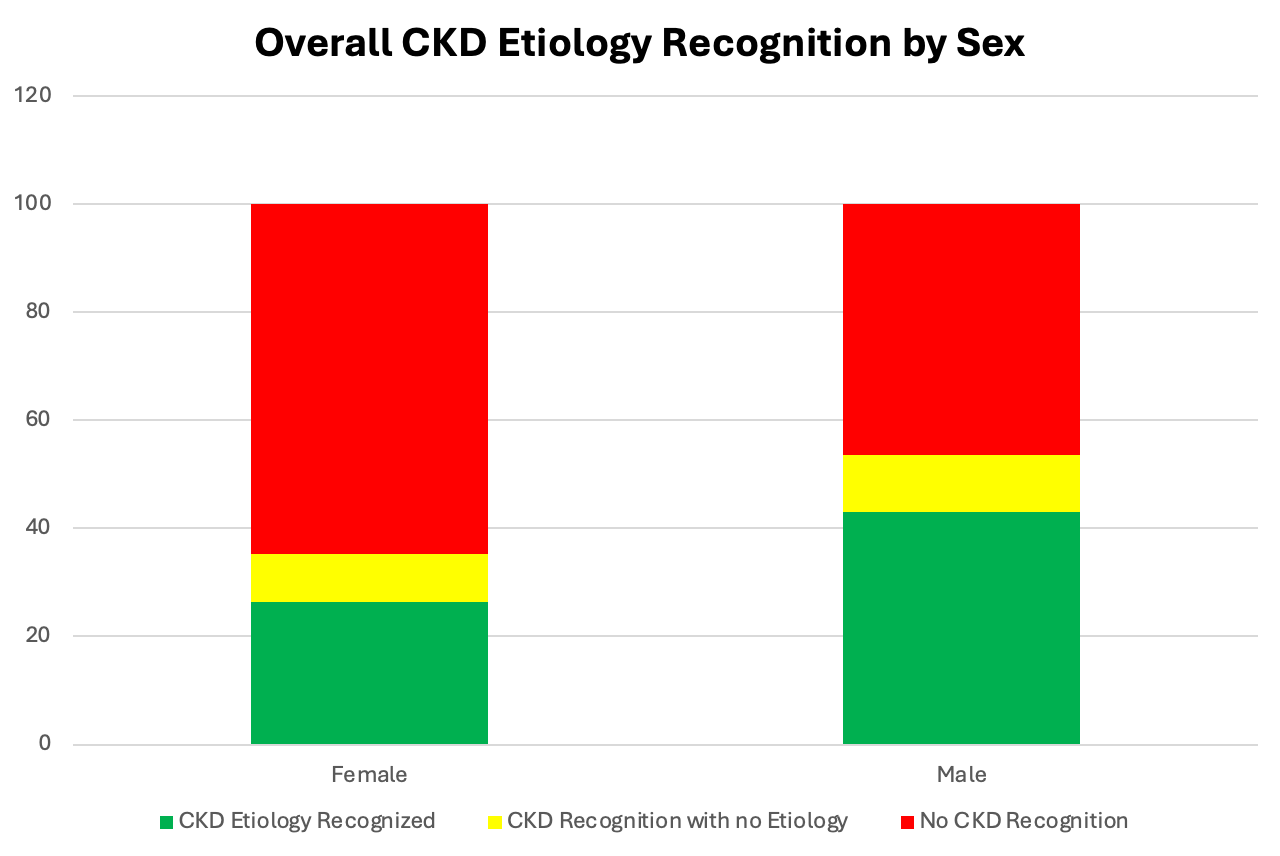
